# Meta-Analysis of Overall Survival in Intramedullary Spinal Gliomas: Comparing Gross Total Resection to Subtotal Resection and Biopsy

**DOI:** 10.64898/2026.03.11.26348187

**Authors:** Mohammad Hamo, Matthew Jarrell, James Shi, Carl Townsend, Yifei Sun, Travis Atchley, Nicholas M.B. Laskay, Dagoberto Estevez-Ordonez

## Abstract

**Background and Objectives:** Intramedullary spinal cord tumors (IMSCTs) are rare, and the extent of surgical resection may influence overall survival (OS). Gross total resection (GTR) may offer superior outcomes compared to subtotal resection (STR) or biopsy. Our study seeks to quantify the benefits of resection extent on OS in patients with spinal gliomas (SGs).

**Methods:** A systematic review was conducted using the following databases: Scopus, Embase, and PubMed. Studies reporting OS in patients who underwent GTR, STR, or biopsy for low- or high-grade SG. We used a random-effects model to calculate pooled hazard ratios (HRs) and 95% confidence intervals (CIs); this was performed separately for low-grade (WHO grade I-II) and high-grade (III-IV) SGs. Subgroup analysis was performed for radiotherapy. I^2^ statistic and Cochran’s Q tests evaluated study heterogeneity, Egger’s and funnel plot asymmetry tests assessed publication bias, and Risk Of Bias In Non-randomized Studies of Exposure (ROBINS-E) evaluated individual study bias.

**Results:** In a pooled analysis of 5 studies, GTR was not associated with improvement in OS compared to STR or biopsy in high grade SGs (HR=0.48, 95% CI: 0.19 –1.26). However, low-grade SGs revealed significant benefit in overall survival with GTR (HR=0.27, 95% CI: 0.15–0.46). Patients treated with radiotherapy were associated with worse outcomes following GTR in low-grade SGs (HR=1.48, 95% CI: 1.30–1.69) but no survival differences in high-grade SGs (HR=1.21, 95% CI: 0.52–2.83). ROBINS-E determined only 1 study with high risk of bias.

**Conclusion:** GTR for intramedullary spinal gliomas may not confer a significant benefit in overall survival for high-grade lesions but may provide benefit in lower grades. Radiotherapy confers a worse survival in lower-grade tumors, potentially due to their infiltrative nature. Future studies should stratify outcomes based on tumor biology, as well as follow functional outcomes overtime.

## Introduction

Intramedullary spinal cord tumors (IMSCTs) consist of 20-30% of primary spinal cord tumors.^1^ Around 80% of intramedullary tumors consist of spinal gliomas (SG), and outcome predictors include tumor biology, impact on function, and surgery outcome.^2^ SG differ in histologic grade and invasiveness, with glioblastoma being rarer (1%) and particular invasive.^3^ Surgical treatment options often involve gross total resection, but glioma infiltration and ill-defined margins increase the risk for functional decline and adverse outcomes.^4^ Lower-grade SGs can present with dissection planes that allow gross total resection (GTR),^5^ but the impact of GTR compared to other surgical methods is ill-defined.

Previous work underscores the need to balance GTR benefits against the risk of post-operative neurological decline.^6^ This balance is particularly important when considering modes of treatment for SGs that differ in tumor biology and infiltration.

We performed a meta-analysis for studies evaluating the survival benefit of GTR for treatment of low- and high-grade intramedullary spinal cord gliomas. The primary outcome was overall survival in GTR compared to subtotal resection (STR) or biopsy. We also sought to compare functional outcomes, adjuvant radiotherapy, and other relevant post-operative outcomes.

## Methods

### Search Strategy and Information Sources

This study was registered in the International Platform of Registered Systematic Review and Meta-analysis Protocols (INPLASY ID: INPLASY202340085). This manuscript complies with the Preferred Reporting Items for Systematic reviews and Meta-Analyses (PRISMA) guidelines.^7^ Databases, including Embase, Scopus, and PubMed, were queried in December 2025 for peer-reviewed studies involving surgical resection of spinal gliomas. Our search strategy targeted adult populations (>18 years), with no study date restrictions.

### Eligibility Criteria

We included studies that reported surgical outcomes in patients aged 18 years or older undergoing resection of either low- or high-grade intramedullary spinal cord gliomas. Studies were excluded if they consisted of case reports, conference abstracts, reviews, non-English publications, patients with prior intramedullary surgery, extramedullary tumors, or lacked detailed follow-up data.

### Data Collection and Extraction

Two reviewers (MJ and MH) independently screened studies for eligibility using Covidence, with discrepancies resolved with consensus or a third reviewer (DEO). Duplicates were automatically removed. Full text reviews were conducted by two reviewers using a shared spreadsheet, and collected data included demographic information, tumor and resection type, and radiation therapy.

### Outcomes and Statistical Analysis

We separated analyses based on World Health Organization (WHO) tumor grade classification, with low-grade (I and II) and high-grade (III and IV) spinal cord gliomas.^8^ The primary outcome was overall survival comparing gross total resection (GTR) to subtotal resection (STR) or biopsy.

Secondary outcomes included post-operative functional outcomes recorded with Modified McCormick Scale (MMCS), as well as impact of radiotherapy on survival. A random-effects model calculated hazard ratios (HR) with 95% confidence intervals (CI). Available data analysis with no imputations for missing data was performed. Bias assessment was done using the Egger’s test with funnel plot method to assess publication bias. The Risk Of Bias In Non-randomized Studies of Exposure (ROBINS-E) evaluated study bias, with domains involving exposure measurements and participant selection. The Grading of Recommendations Assessment, Development and Evaluation (GRADE) assessment was applied to both GTR and radiotherapy in treating both grades of spinal gliomas.^9^ Leave-one-out sensitivity analysis was conducted to evaluate the influence of individual studies on pooled effect estimates.

## Results

### Study Selection and Bias Assessment

We screened 1183 total studies, with 241 removed automatically as duplicates. Full-text reviews were done for 192 studies, with 5 studies included for data extraction.^10–14^ This study follows the Preferred Reporting Items for Systematic Reviews and Meta-Analyses (PRISMA), with a flow diagram displayed in Fig. 1.^7^

**Fig 1.**
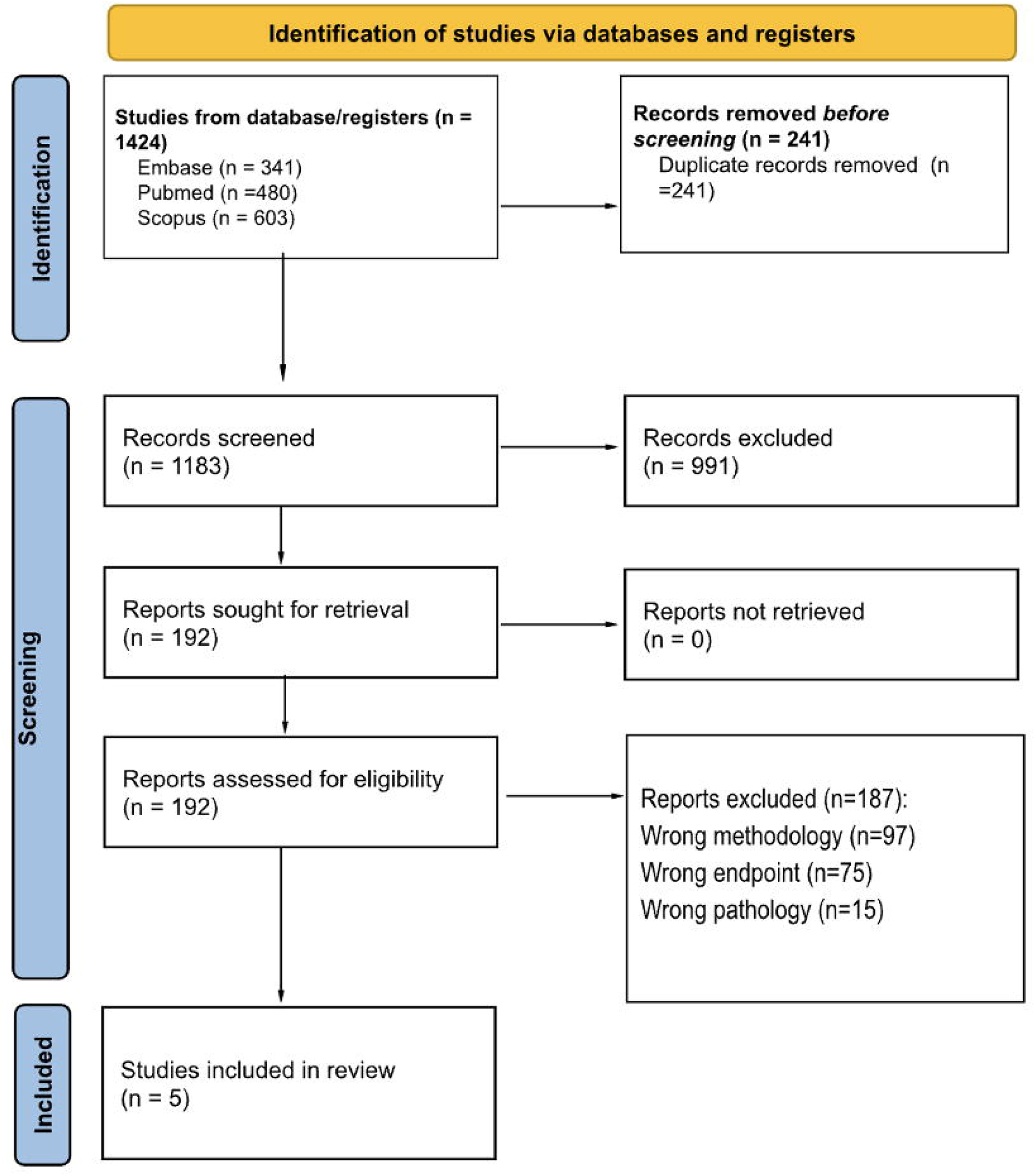
PRISMA flow diagram

Egger’s test assessing publication bias revealed no significant differences between studies (p=0.543) (Supplementary Digital Content, Figure S1). The ROBINS-E tool overall reported 4 studies with some concern and 1 study with high concern for bias. Specifically, most studies carried high concern for bias in selecting participants for their study or analysis. ROBINS-E tool summaries can be found in Supplementary Digital Content, Figure S2. The GRADE assessment reported moderate certainty in recommending GTR and radiotherapy in both low- and high-grade SGs (Supplementary Digital Content, Figure S3,S4). Leave-one-out sensitivity analysis revealed Chalif et al. WHO grade III findings hurt survival in GTR for high-grade SGs, and its removal conferred significant survival benefit in GTR (Supplementary Digital Content, Table S1). Sensitivity analysis for low-grade lesions revealed removal of either Zhang et al. or Diaz-Aguilar et al. removed significant survival benefit of GTR (Supplementary Digital Content, Table S2).

### Study Characteristics

The included studies comprised 1477 patients. Among these, 205 patients underwent GTR, while 749 underwent STR, biopsy, or no surgery. The remainder of the patients did not have a reported extent of surgery. Tumor types included glioblastoma (GBM), anaplastic astrocytoma (AA), diffuse astrocytoma (DA), pilocytic astrocytoma (PA), astrocytoma not-otherwise-specified (A-NOS), glioma not-otherwise-specified (G-NOS), low-grade astrocytoma, and high-grade astrocytoma. GBM and high-grade astrocytoma were the most reported types. Resection types included GTR, STR, and biopsy. Adjuvant therapy varied, with only McGirt et al not reporting postoperative radiotherapy. MMCS scores were only reported in Zhang et al and McGirt et al studies. Patient demographics and other management information can be found in Table 1.

**Table 1:** Characteristics of Included Studies and Cohorts.

| Table 1: Characteristics of Included Studies and Cohorts |  |  |  |  |  |  |  |  |
| --- | --- | --- | --- | --- | --- | --- | --- | --- |
|  | Ryu et al. |  | Zhang et al. |  | McGirt et al. |  | Diaz-Aguilar et al.* | Chalif et al.* |
| Tumor Type | Low Grade A | High Grade A | High Grade A | Low Grade A | High Grade AA | High Grade GBM | DA, PA, A-NOS, G-NOS | WHO Grade IV (non-GBM), GBM, WHO III, WHO II |
| Gender (M: F) | 10:4 | 7:5 | 29:25 | 38:16 | 11:16 | 4:4 | 315:246 | 406:341 |
| Mean Age (years) | 34.2 (14-57) | 47.5 (19-76) | 35.5 (8-63) | 27.0 (4-54) | 31.3 (1.5-61) | 20.5 (7-51) | 28 (1-94) | Median 41 (IQR 17-58) |
| Resection Types | GTR | GTR, STR | GTR, STR, Biopsy | GTR, STR, Biopsy | GTR, STR | STR | GTR, STR, No surgery | GTR, STR, Biopsy |
| Gross Total Resection (N) | 14 | 7 | 23 | 35 | 12 | 0 | 87 | 27 |
| Radiation Therapy (No: Yes) | 11:3 | 5:7 | 14:30 | 16:32 | 2:25 | 0:8 | 323:220 | 290:442 |
| Chemotherapy (No: Yes) | 13:1 | 10:2 | 25:19 | 40:8 | 14:13 | 1:7 | NR | 394:324 |

A-NOS = Astrocytoma not otherwise specified, DA = Diffuse Astrocytoma, G-NOS = Glioma not otherwise specified, GBM = Glioblastoma, AA = Anaplastic Astrocytoma, PA = Pilocytic Astrocytoma, Non-Pilocytic Astrocytoma = NPA, A = Astrocytoma, NR = Not Reported
\*Study did not report individual characteristics for each grade, so overall cohort characteristics were collected

### Survival and Neurologic Outcomes

Patients undergoing GTR did not demonstrate improved overall survival compared to those who underwent STR or biopsy for high grade SGs (Fig. 2A; HR=0.48, 95% CI: 0.19 –1.26). However, GTR for low-grade SGs revealed significant benefit in overall survival (Fig. 2B; HR=0.27, 95% CI: 0.15–0.46).

**Fig 2.**
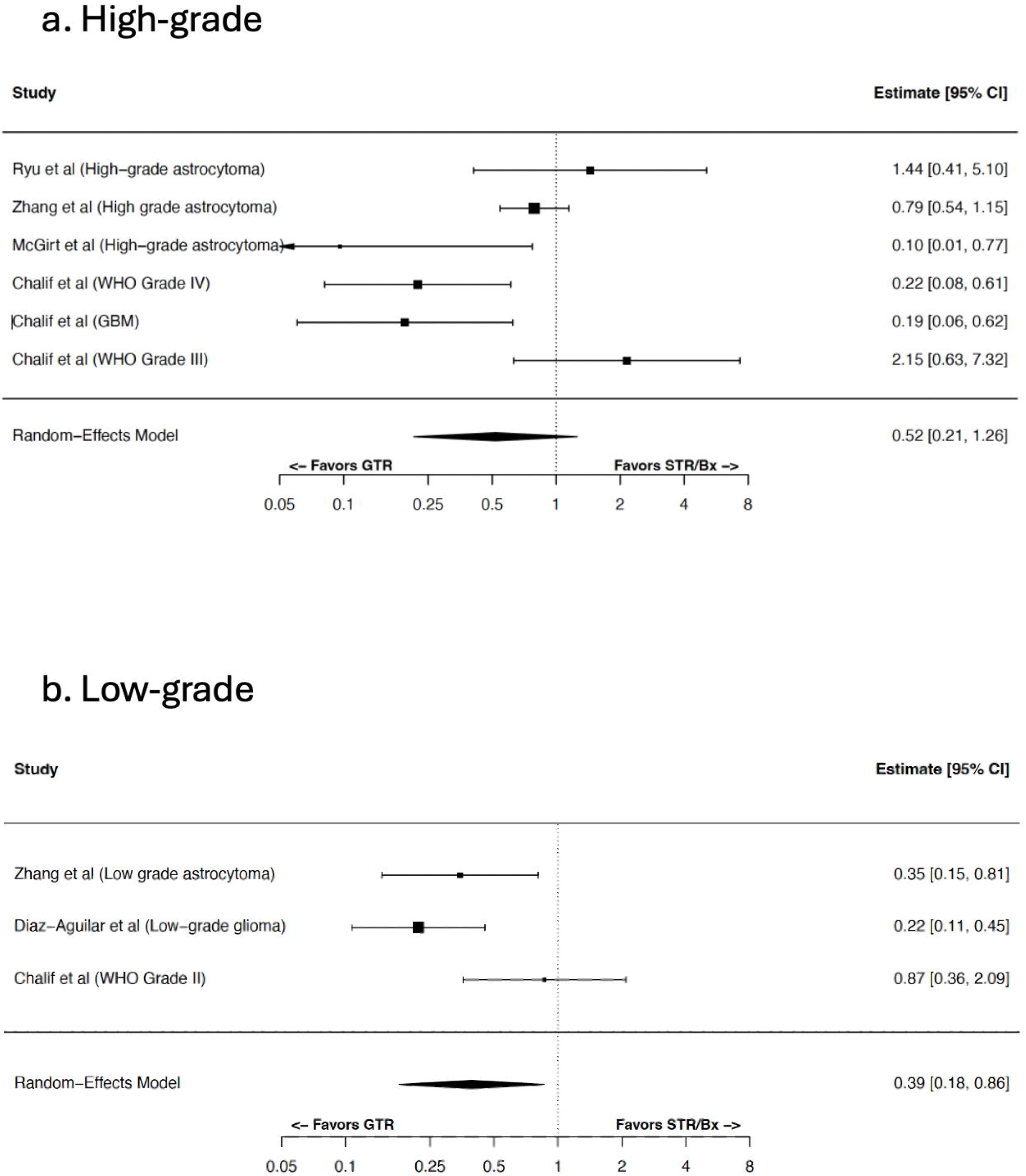
Forest plot comparing overall survival in gross total resection versus subtotal resection or biopsy in high- and low-grade spinal gliomas This forest plot depicts overall survival in high-grade spinal cord lesions comparing gross total resection (GTR) with subtotal resection (STR) or biopsy (Bx) including hazard ratios and 95% confidence intervals (CI). Pooled HR and 95% CI are generated from random-effects model.

Among individual studies, Zhang et al reported outcomes in 23 subjects with high-grade and 35 with low-grade astrocytoma; there was a significant survival benefit in GTR of low-grade astrocytoma (HR=0.346, 95% CI: 0.148-0.806). Diaz-Aguilar et al reported survival benefits in GTR of low grade, WHO I-II SGs (diffuse, pilocytic, and gliomas not-otherwise specific) (HR=0.22, 95% CI: 0.13–0.55). Zhang et al and McGirt et al reported MMCS, which mildly increased comparing pre- and post-operative outcomes. Pooled analysis for post-operative radiotherapy in low-grade tumors was associated with worse overall survival (Fig. 3b; HR=1.48, 95% CI: 1.30–1.69), but was not associated with survival differences in high-grade tumors (Fig. 3a; HR=1.21, 95% CI: 0.52–2.83).

**Fig 3.**
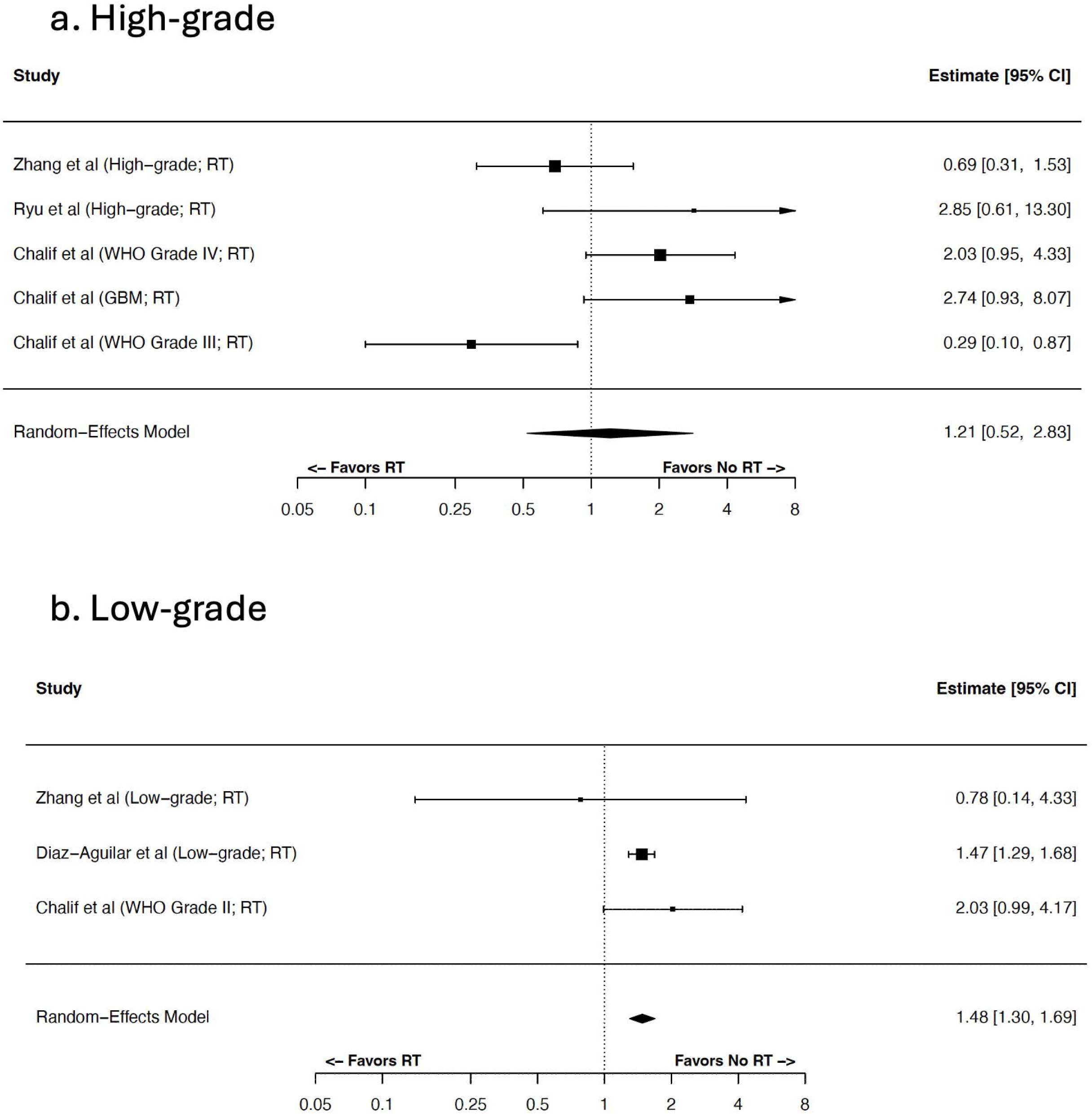
Forest plot comparing overall survival with radiotherapy treatment in high- and low-grade spinal cord gliomas RT = Radiotherapy

This forest plot depicts overall survival in high-grade spinal cord tumors comparing treatment with radiotherapy including hazard ratios (HR) and 95% confidence intervals (CI). Pooled HR and 95% CI are generated from random-effects model.

A summary of primary and secondary outcomes can be found in Table 2.

**Table 2:**
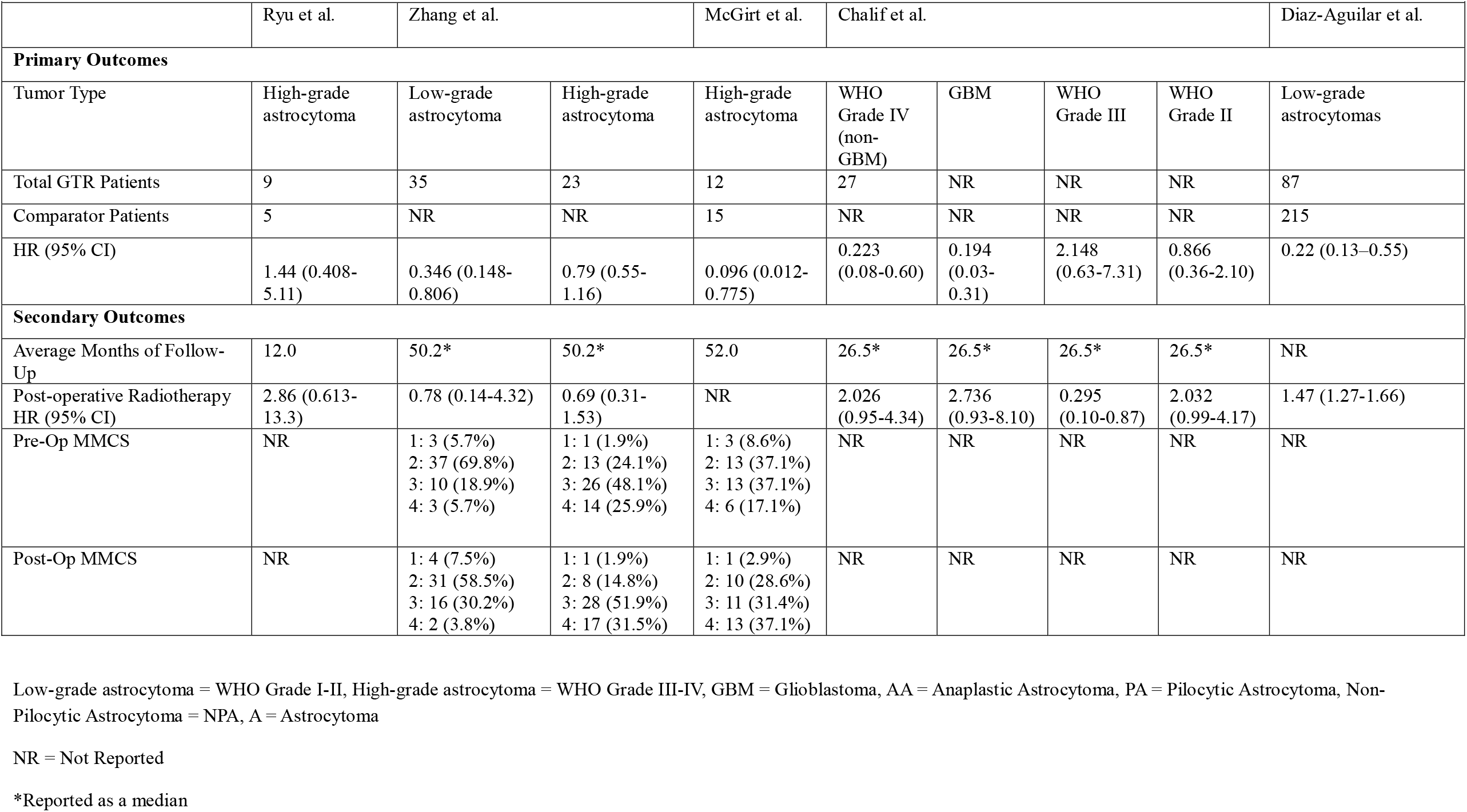
Primary and Secondary Outcomes of Included Studies.

## Discussion

This meta-analysis included 5 studies comparing GTR surgical management of either low-grade or high-grade SGs; low-grade SGs were associated with improved survival with GTR, while high-grade SGs found no significant survival benefit with GTR. Future, multi-center studies could help elucidate benefits of GTR by studying individual tumor histology and grade, as well as longitudinal functional outcomes. Post-operative radiotherapy was associated with worse survival in low-grade spinal gliomas, but no survival differences in high-grade tumors. Sensitivity analysis revealed inconsistencies in survival benefit of GTR in low-grade SGs, highlighting the importance of future, multi-intuitional studies that can elucidate the benefits in GTR stratified by tumor grade.

Gross total resection remains an ideal treatment goal, but tumor grade and infiltration can limit resection extent. 40% of IMSCTs consist of astrocytomas, 25% being high-grade.^15,16^ Higher-grade astrocytomas can be more infiltrative, complicating surgical approaches and limiting extent of resection.^5^ Both Diaz-Aguilar et al and Zhang et al report improved survival in GTR for lower-grade astrocytomas, potentially attributed to lower-infiltrative nature and tumor resection planes.^10,14^ Glioblastoma comprises 7.5% of intramedullary spinal cord tumors, and its rarity poses challenges to standardizing management protocols.^17^ Additionally, its infiltrative nature could result in severe post-operative functional impairment; due to its aggressiveness, a study has recorded a median time to death of 20 months post-resection.^18^ However, McGirt et al stratifies survival by tumor type, reporting improved survival with GTR in spinal cord GBM.^13^ Future studies stratifying GTR treatment based on tumor grade and histology could better determine survival outcomes with resection extent.

A concern for GTR involves impact on neurologic function. A clinical trial assessing outcomes from resection in intramedullary spinal cord tumors reported 60% of patients experienced deterioration in upper and lower extremity function, regardless of resection extent.^19^ Additionally, another study found that 28% of patients in a 302 patient cohort experienced complications post-operatively, with only readmission being significantly associated with astrocytomas compared to ependymoma and hemangioblastoma.^5^ However, studies have evaluated methods to improve functional outcomes. The use of intraoperative neurophysiological monitoring has shown to improve functional outcomes and McCormick scores post-operatively.^20^ Additionally, the use of distal muscle-recorded transcranial motor evoked potentials during operations has shown to predict post-operative motor outcomes with tumor resection.^21^ Zhang et al and McGirt et al were the only included studies to report functional outcomes, with both reporting moderate functional impairments post-operatively through increasing MMCS scores. Future studies should longitudinally evaluate functional outcomes across resection extent and tumor biology, while integrating intraoperative monitoring.

Benefits from adjuvant radiotherapy are conflicting and vary regardless of resection extent. Studies report radiotherapy was associated with decreased mortality in high-grade lesions, but increased mortality in low-grade lesions; however, treatment was associated with reduced time to recurrence.^22^ Our meta-analysis suggests that radiotherapy may be associated with worsening survival in patients with low-grade tumors but no survival differences in high-grade lesions. Additionally, previous studies find infiltrative tumors gain improved survival with radiotherapy, causing radiotherapy to be linked to more aggressive disease and poorer outcomes.^23^ In low-grade tumors, radiotherapy is used for more infiltrative disease or cases with subtotal resection, potentially biasing survival outcomes. In contrast, higher-grade lesions traditionally receive radiotherapy due to their infiltrative nature. Stratified cohorts analyzing similar tumor grades and infiltration could better evaluate the survival benefit of radiotherapy.

## Limitations

One of the limitations is the number of studies included. Studies were only included reporting hazard ratios and confidence intervals with total resection of intramedullary SGs. Additionally, all included studies were retrospective, increasing risk for selection bias and confounding. Another aspect was the heterogeneity of tumor types and infiltration. Future studies would benefit from stratified results separated by tumor biology. Older studies also do not include molecular differentiation of tumor subtypes, which limits stratification from current standard practice. Studies also may vary in their classification of total resection, which could cause misclassification bias. Future studies should introduce a standardized approach to defining resection extent. Functional outcomes are an essential aspect of post-operative care and should be included in studies. Sparse reporting of neurologic function could hinder the comprehensive evaluation of resection extent in spinal gliomas.

## Conclusion

This meta-analysis suggests GTR may not significantly improve survival outcomes compared to STR or biopsy for high-grade intramedullary spinal gliomas, but low-grade lesions may benefit from more aggressive surgical management. However, tumor biology and invasion have been shown to influence outcomes. Few studies reported functional outcomes, with patients showing moderate worsening post-operatively. Additionally, adjuvant radiotherapy was associated with worse survival in low-grade tumors, but this may result from its need in more infiltrative tumors. Future studies would benefit from stratified analysis of resection types based on tumor biology, including molecular makeup, and invasion.

## Data Availability

All data used in the present study can be acquired on authors request.

## Acknowledgements

The authors have no additional acknowledgements to declare.

## Abbreviations

IMSCTs: Intramedullary spinal cord tumors
MMCS: Modified McCormick Scale
PRISMA: Preferred Reporting Items for Systematic Reviews and Meta-Analyses
REM: Random Effects Model
ROBINS-E: Risk Of Bias In Non-randomized Studies -of Exposure

