## Supplementary Materials for "Meta-Analysis of Overall Survival in Intramedullary Spinal Gliomas: Comparing Gross Total Resection to Subtotal Resection and Biopsy"

**Supplementary Digital Content**

Figure S1: Funnel plot for detection of publication bias

Figure S2: Risk of Bias in Non-randomized Studies of Exposure (ROBINS-E) tool

Figure S3: Grading of Recommendations Assessment, Development and Evaluation (GRADE) assessment for recommending gross total resection (GTR) in high- and low-grade spinal gliomas

Table S1. Leave-one-out sensitivity analysis for high-grade spinal gliomas

Table S2. Leave-one-out sensitivity analysis for low-grade spinal gliomas

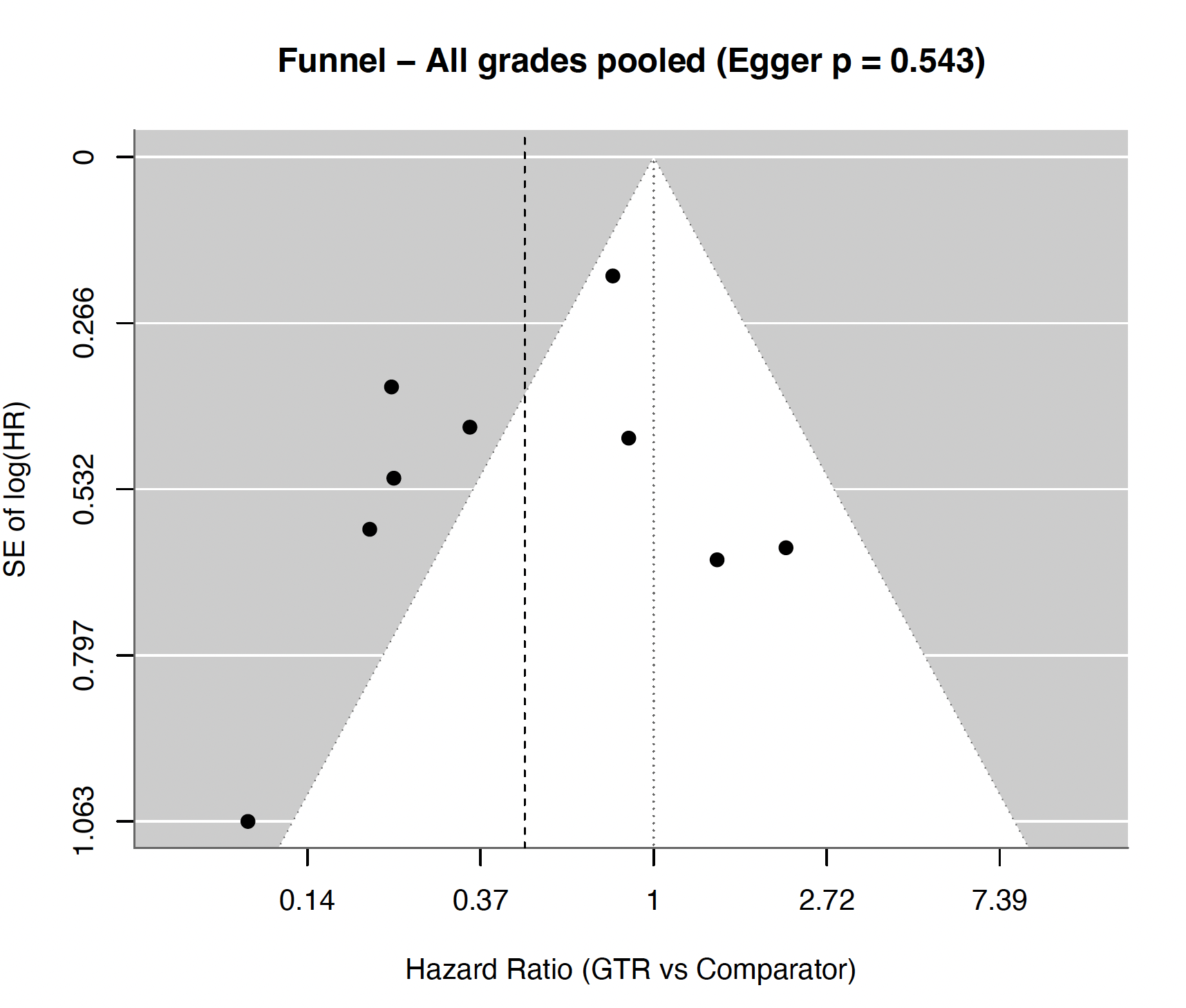

Figure S1: Funnel plot for detection of publication bias

Distribution may not reflect bias in included studies (p=0.543).

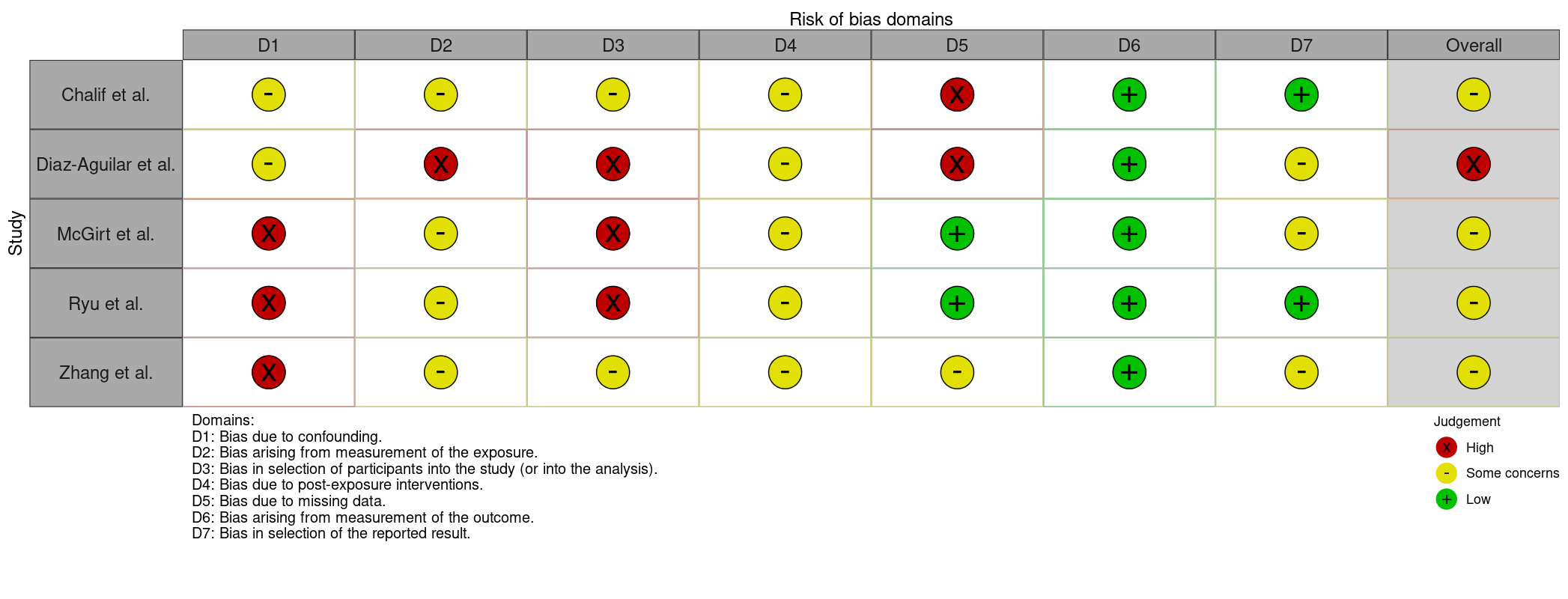

Figure S2: Risk of Bias in Non-randomized Studies of Exposure (ROBINS-E) tool

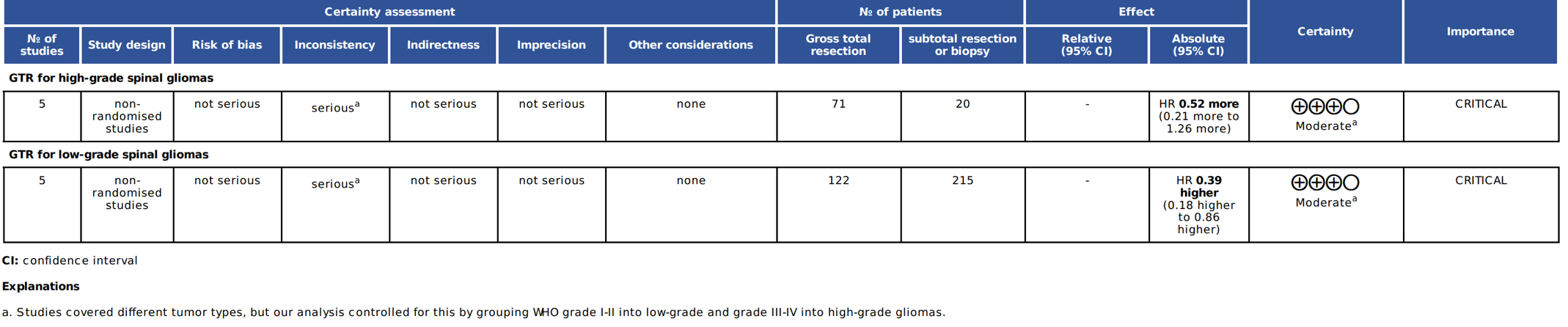

Figure S3. Grading of Recommendations Assessment, Development and Evaluation (GRADE) assessment for recommending gross total resection (GTR) in high- and low-grade spinal gliomas

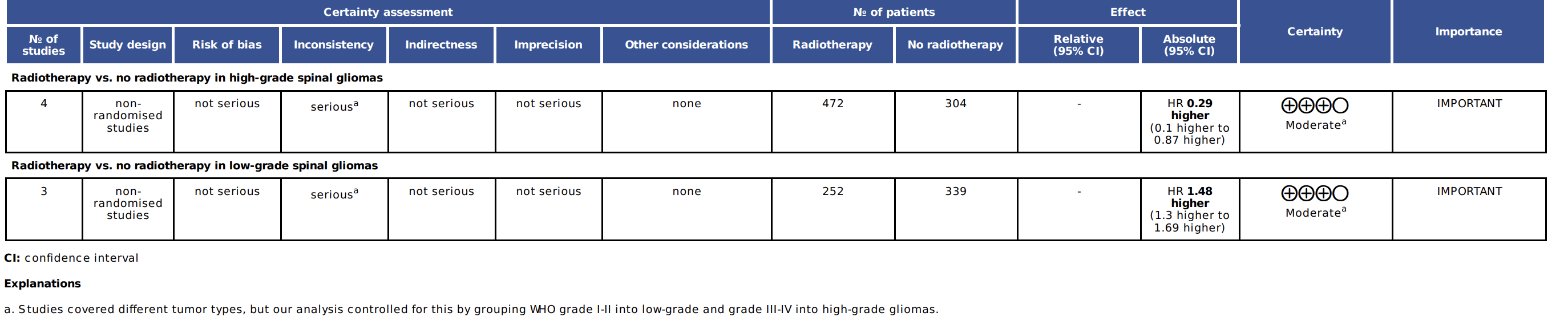

Figure S4. Grading of Recommendations Assessment, Development and Evaluation (GRADE) assessment for recommending radiotherapy in high- and low-grade spinal gliomas

| **Study** | **Estimate** | **SE** | **Z** | **P** | **Lower95** | **Upper95** | **Q** | **Q_p** | **tau2** | **I2** | **H2** |
| --- | --- | --- | --- | --- | --- | --- | --- | --- | --- | --- | --- |
| **Ryu et al (High-grade astrocytoma)** | 0.424084040961774 | 0.509501752880895 | -1.68365197710355 | 0.0922489473200848 | 0.156229574941883 | 1.15117303407738 | 16.3770687797222 | 0.00255276563441796 | 0.942904202609997 | 79.66752320532 | 4.91823996701498 |
| **Zhang et al (High grade astrocytoma)** | 0.452479065042242 | 0.584292394337653 | -1.35722078482878 | 0.174711068432985 | 0.143962010331421 | 1.42216202614959 | 15.4739722142959 | 0.0038126880413247 | 1.24095440120906 | 74.8827488167439 | 3.98132738612189 |
| **McGirt et al (High-grade astrocytoma)** | 0.627984888127781 | 0.454240191451769 | -1.02421402828169 | 0.305734194206749 | 0.257809692587421 | 1.5296749154733 | 14.4702476009726 | 0.00593602593800498 | 0.760455353115493 | 78.1682692442639 | 4.58048888193283 |
| **Chalif et al (WHO Grade IV)** | 0.622166654917667 | 0.51056594042024 | -0.929453476252298 | 0.352654125133669 | 0.228724251448441 | 1.69239310672177 | 12.826340725494 | 0.01215624601818 | 0.917804148600484 | 77.1087068175955 | 4.36847316589634 |
| **Chalif et al (GBM)** | 0.632790576305558 | 0.490938562550492 | -0.932124281132909 | 0.351272300920711 | 0.241753260922495 | 1.65633303945172 | 13.2294600745269 | 0.0102073676277838 | 0.843870742780829 | 77.1864257933659 | 4.38335523816871 |
| **Chalif et al (WHO Grade III)** | 0.402263553200787 | 0.447605759589339 | -2.03448633242918 | 0.0419025852518985 | 0.167304634729331 | 0.967193565770078 | 14.1764755628108 | 0.00675260514711363 | 0.6601252690196 | 73.0291107492101 | 3.70770125783195 |

Table S1. Leave-one-out sensitivity analysis for high-grade spinal gliomas

| **Study** | **Estimate** | **SE** | **Z** | **P** | **Lower95** | **Upper95** | **Q** | **Q_p** | **tau2** | **I2** | **H2** |
| --- | --- | --- | --- | --- | --- | --- | --- | --- | --- | --- | --- |
| **Zhang et al (Low grade astrocytoma)** | 0.425941889291749 | 0.684692154009523 | -1.2464760215711 | 0.212589686969362 | 0.111311320477299 | 1.62990154348609 | 5.55839196138856 | 0.0183923006365208 | 0.76990437937482 | 82.0091852653337 | 5.55839196138857 |
| **Diaz-Aguilar et al (Low-grade glioma)** | 0.542795726966482 | 0.458645623042228 | -1.33223166767764 | 0.182784073446341 | 0.220920774957979 | 1.33363284312719 | 2.16185384266084 | 0.141473915560637 | 0.226181105990264 | 53.7434039125797 | 2.16185384266084 |
| **Chalif et al (WHO Grade II)** | 0.266087510798455 | 0.280217164484896 | -4.72465717356995 | 2.30503880685515E-06 | 0.153639614198631 | 0.460835337111574 | 0.636110970438098 | 0.425122589716894 | 0 | 0 | 1 |

Table S2. Leave-one-out sensitivity analysis for low-grade spinal gliomas
